# Barriers and Enablers to Scaling the AURUM Management Development Programme: District Manager Perspectives from the Western Cape, South Africa

**DOI:** 10.64898/2026.05.28.26354359

**Authors:** Constance Mongwenyana-Makhutle, Aneesa Moolla, Danleen Hongoro, Tembeka Sineke, Khumbo Shumba, Jacqui Miot, Dorina Onoya

## Abstract

**Background:** Strong management capacity is essential for effective primary healthcare (PHC) service delivery and health system strengthening [1]. The AURUM Management Development Programme (MDP) was implemented to strengthen district and PHC leadership in the Western Cape province of South Africa. This study explored the contextual barriers and enabling conditions influencing the scalability of the programme within district health systems.

**Methods:** This study employed a qualitative exploratory design to investigate barriers and enablers associated with scaling the MDP. In-depth interviews were conducted with purposively selected district health managers from three Western Cape districts. Interviews were audio-recorded, transcribed verbatim, and analysed thematically using NVivo 14. The study explored perceptions regarding programme adaptability, district readiness, implementation challenges, and enabling conditions for sustainability and scale-up.

**Results:** Twenty participants (7 males and 13 females) from the Cape Winelands, Garden Route, and Cape Town Metro district health offices were interviewed. The MDP was viewed as relevant, practical, and adaptable to district health system contexts. District readiness for implementation emerged as an important determinant of perceived programme success. High readiness was characterised by clear team roles, strong management structures, decentralised decision-making, digital tool utilisation, ongoing mentorship systems, and prior exposure to PHC reforms such as the Ideal Clinic Realisation and Maintenance (ICRM) programme. Lower readiness was associated with staff shortages, operational pressures, limited leadership support, and partially functional health systems. Key enabling factors included integration with existing training structures, visible improvements in service delivery, mentorship support, and active engagement from district leadership.

**Conclusion:** The MDP demonstrates potential for scalability within South Africa’s public health system. However, successful scale-up depends on district-level readiness, supportive leadership structures, integration into existing training and management systems, and sustained mentorship and implementation support.

## Introduction

Effective leadership development within health systems is increasingly recognised as essential for strengthening primary health care (PHC) and supporting health sector reforms, particularly in low- and middle-income countries (LMICs) [1–4]. In South Africa (SA), ongoing decentralisation of health services and the implementation of Universal Health Coverage (UHC) through initiatives such as the National Health Insurance (NHI) underscore the importance of capacitated district management teams [5]. The AURUM Management Development Programme (MDP) was implemented to equip middle and senior health managers with the leadership and operational skills required to navigate complex health system reforms and improve service delivery.

Improving management capacity in PHC is recognised globally as a critical strategy for enhancing health system performance [6, 7]. However, PHC managers in SA often operate within complex environments characterised by workforce shortages, fragmented services, resource constraints, and limited formal leadership training [3, 8–12]. Previous initiatives in the Western Cape, including bottom-up district management innovations and the Appointment System Learnings Initiative (ASLI), demonstrated the value of distributed leadership, organisational learning, and adaptive management approaches in strengthening health system resilience [5, 13].

Despite these advances, limited evidence exists regarding the conditions required to successfully scale leadership development programmes within district health systems. Sustainable scale-up requires alignment with existing organisational structures, supportive supervision systems, and broader health system priorities [13–15]. Middle-level managers are uniquely positioned to bridge top-down policy directives and frontline implementation, provided they are supported with appropriate leadership training and institutional support[9, 11].

This study explores the perspectives of district managers from three districts in the Western Cape regarding the scalability of the AURUM MDP. Through qualitative analysis, the study identifies the contextual barriers and enabling conditions influencing the broader rollout and sustainability of leadership development initiatives within district health systems in South Africa

## Methodology

### Study design and setting

This study employed a qualitative exploratory design to examine barriers, enablers, and perceived feasibility of scaling the AURUM Management Development Programme (MDP) across district health systems in the Western Cape Province of South Africa. The study was conducted across three purposively selected health districts representing diverse geographic and service delivery contexts: Cape Winelands (rural), Garden Route (peri-urban), and Cape Town Metro (urban). District health managers were recruited using snowball sampling based on their involvement in primary health care management and engagement with the MDP implementation process.

### Data collection

This study employed a retrospective qualitative data collection approach. Twenty in-depth interviews were conducted between January and June 2025 with district health managers from three health districts in the Western Cape Province. Participants were selected using snowball sampling based on their direct involvement in primary health care oversight and exposure to the MDP. All interviews were conducted virtually in English and lasted between 30 and 60 minutes. All the written consent forms were signed before the interviews.

### Data analysis

All interviews were audio-recorded and transcribed verbatim in English. To ensure accuracy and intercoder reliability, a second researcher cross-checked the transcripts. To enhance the rigor of the analysis, a subset of two transcripts was independently coded by four researchers. They subsequently convened to reconcile discrepancies and build a consensus-based approach to coding through an iterative process.

A rigorous inductive thematic analysis, informed by principles of grounded theory, was applied to the data. Emergent themes were derived directly from the participant narratives rather than from a pre-defined conceptual framework. As new patterns were identified, additional codes were created, and earlier transcripts were revisited to ensure consistency. A structured codebook was then developed from these themes and aligned with the study’s conceptual framework. All qualitative data were managed using NVivo 14 software. Data saturation was considered achieved when no new themes emerged from the final interviews. The key dimensions for analysis included programme adaptability, financial and resource constraints, district-level readiness and support, and operational challenges and enabling factors.

Although a separate economic evaluation of the MDP was conducted within the broader project, this paper does not present formal costing or cost-effectiveness analyses. Instead, district managers’ reflections on budgeting constraints, resource requirements, and financial feasibility are explored qualitatively to understand the programme’s scalability within routine health system structures.

### Ethical approval

To maintain confidentiality and anonymity, all identifiers were removed from the final analytic data in compliance with regulatory requirements and ethical guidelines for research with human subjects in SA and institutional policies (ICH Good Clinical Practice). This study was approved by the Witwatersrand Human Research Ethics Committee (HREC) of the University of the Witwatersrand (ETHICS REFERENCE NO: 240713).

## Results

The stakeholder group comprised 20 key informants representing a diverse cross-section of health system leadership and specialised clinical support across three districts. As detailed in Table 2, participants held strategic and operational roles, including District Directors, Assistant Managers, and leads for specialised units such as Finance and Supply Chain, HIV/AIDS (HAST), and Violence Prevention. Geographically, half of the participants were drawn from the Cape Winelands (n=10), followed by the Cape Town Metro (n=6) and Garden Route (n=4). Demographically, the cohort was predominantly female (65%), though gender distribution varied by district, ranging from a 70% female majority in Cape Winelands to an even gender split in the Garden Route (Table 3). This multidisciplinary composition ensured that the qualitative evaluation captured a broad range of perspectives from both administrative and clinical governance levels.

**Table 2.**
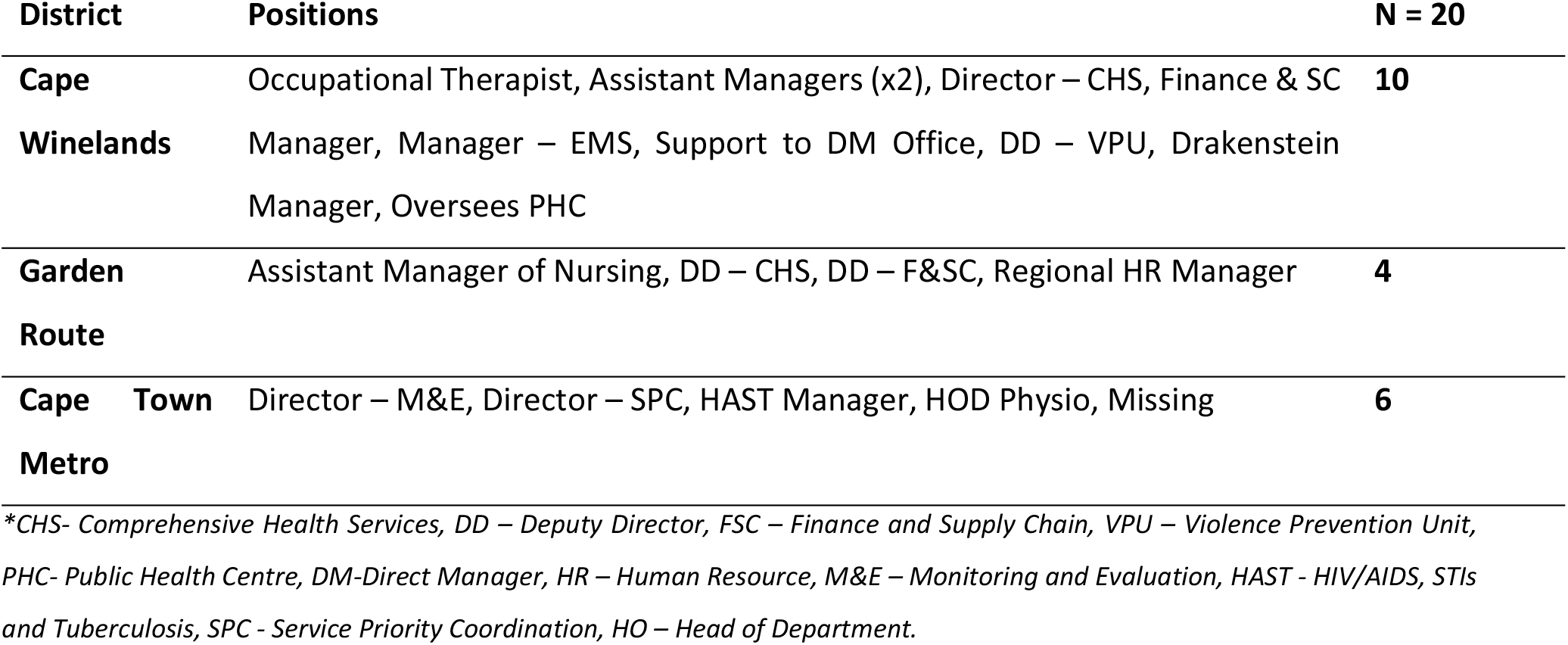
Stakeholder’s Occupations.

**Table 3.** Key Informant Interview Participants’ Demographic Characteristics.

| Gender | Cape Winelands | Garden Route | Cape Town Metro | Total |
| --- | --- | --- | --- | --- |
| **Female** | 7 (70.0%) | 2 (50.0%) | 4(66.7%) | 13(65.0%) |
| **Male** | 3 (30.0%) | 2 (50.0%) | 2 (33.3%) | 7 (35.0%) |
| **Total** | **10(50.0%)** | **4(20.0%)** | **6(30.0%)** | **20 (100%)** |

**Table 4.**
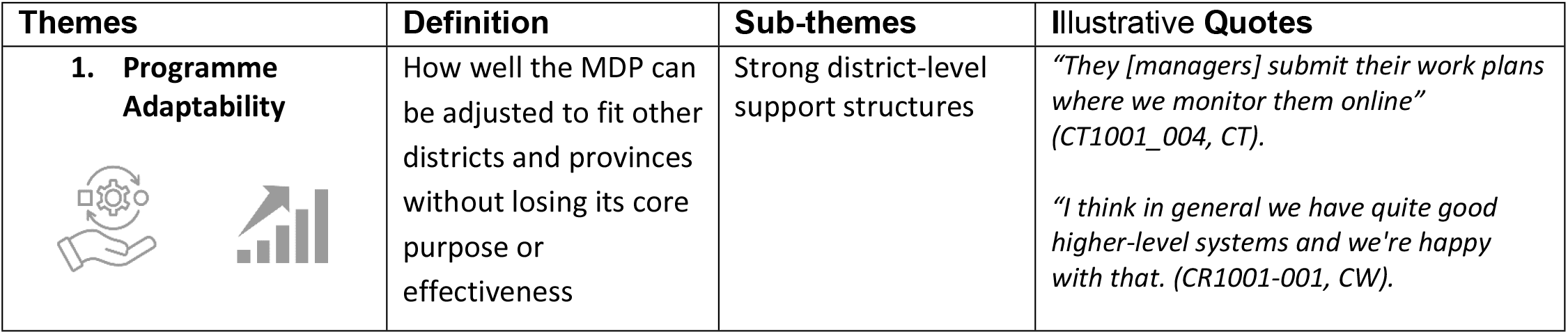
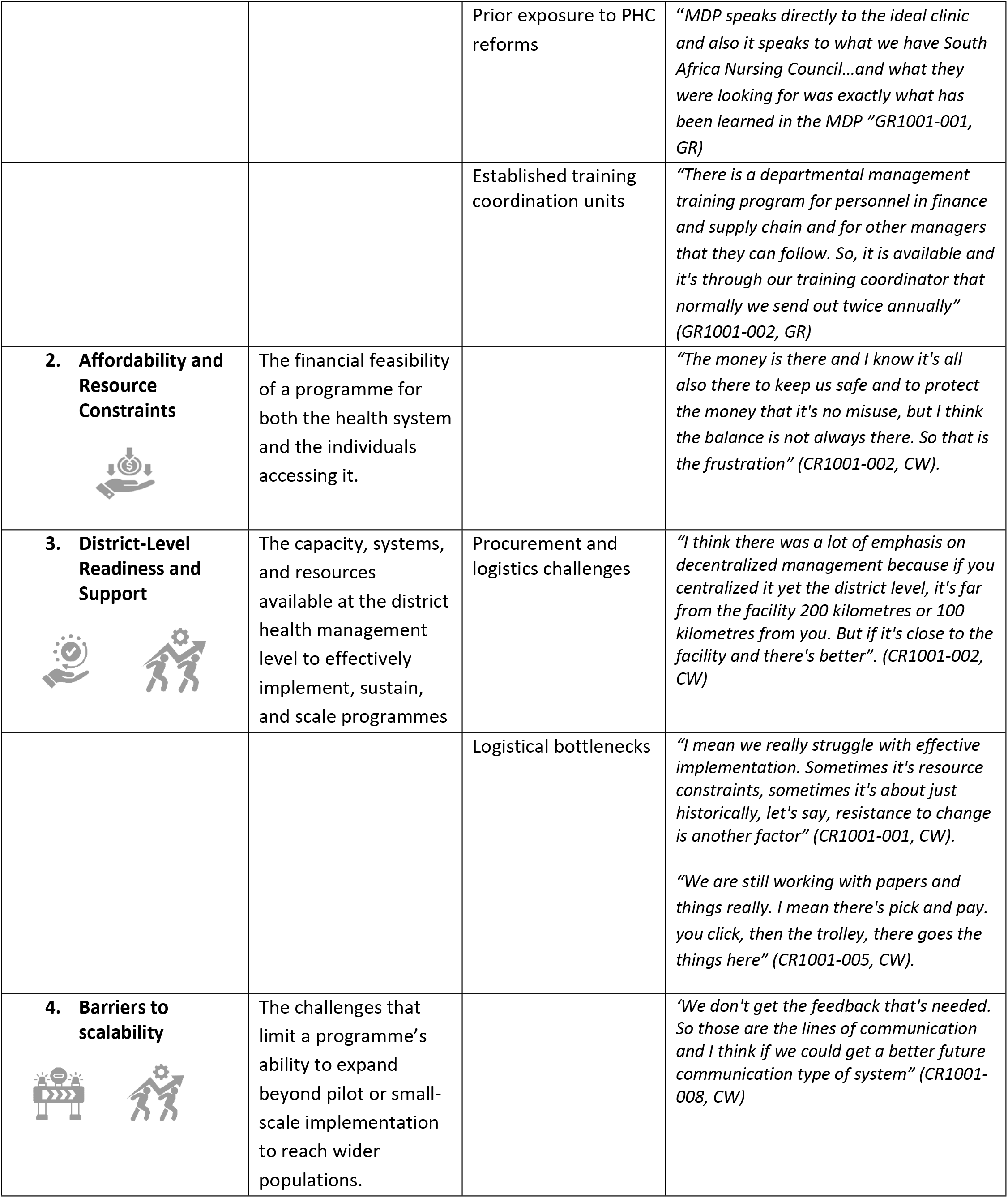
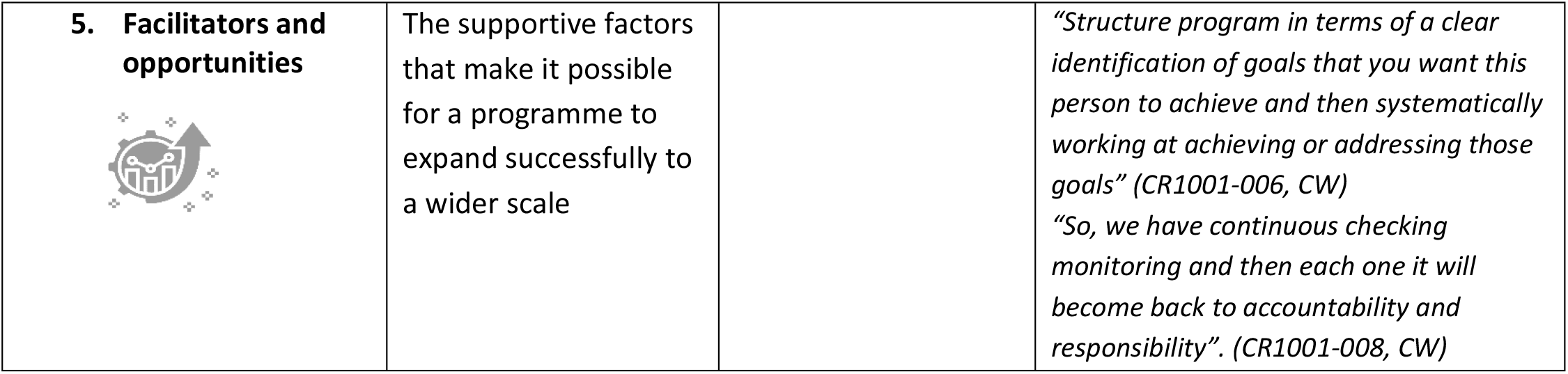
Themes, definitions and quotes.

### 1. Programme Adaptability

Analysis of the qualitative revealed two central findings regarding the scalability of the AURUM MDP. First, the programme’s focus on foundational, universally applicable leadership skills, such as problem-solving and team management, makes it inherently adaptable to diverse healthcare contexts in SA. Second, the effective implementation of the MDP is not uniform and is dependent upon the actual pre-existing operational systems in place as well as the support structures within each district. The results presented in this section detail how districts with robust management frameworks more readily integrated the programme, while those with less strong systems faced distinct barriers to successful scale-up, highlighting the critical role of local context in sustainable expansion.

#### Strong district-level support structures

Some respondents indicated that the strong support provided through the mentorship component of the MDP significantly enhanced their existing work structures. The effective integration of MDP content into their routine operations was reflected in the presence of clearly defined team roles and a flattened management structure with more decentralized responsibilities. This was further enabled by the high levels of professionally qualified staff that were part of the system.

> *“I think it’s working in the sense that our primary healthcare managers are mainly qualified nursing staff. And also, assistant primary care managers. And then we’ve also got family physicians. So, we’re really flattening the structure in the things that we decentralize the responsibilities to the sub-district level. So, I think it works in the sense that the roles are very clear*.*” (CR1001-001, Cape Winelands)*.

Furthermore, the successful integration of the MDP was supported by robust feedback mechanisms, progress tracking, and effective internal communication systems. Formal feedback loops and team engagement were beneficial in providing managers with actionable insights into their leadership styles and programme implementation efforts. The ability to track progress and visualize improvements in key performance indicators enhanced accountability, whilst also providing tangible evidence of the programme’s value, reinforcing its adoption into routine practice. Finally, effective internal communication systems were crucial for building up a shared understanding of programme goals and successes, facilitating knowledge exchange and promoting of continuous improvement across the districts.

> *“So, we have on a regular basis every quarter feedback from our different funded organizations on progress and so I’ve heard the MDP group also presenting information on work that they’ve done to help us improve” (CR1001-001, CW)*.

> *“We have a dashboard which is based on our census figures. So, there are set targets that against which our performance is monitored and it’s the same type of dashboard that we use to manage our own performance and they use the same thing to hold us accountable” (CR1001-006, CW)*.

> *“We use Microsoft Teams in the province for meetings…we also use online platforms for training and our training component has a number of courses which people can take in their own time. They can do it after hours if they can’t do it during the week because the course has been made online. So, we definitely use online platforms” (CR1001-001, CW)*.

Additionally, the district’s existing digital infrastructure and internal support services were found to work seamlessly with the MDP’s materials. This includes the integration of programme-specific policies into workflow. The easily available resource mechanism serves to improve the functioning of higher-level systems.

> *“We’ve got a lot of policies in the Western Cape for the delivery of corporate services as well as strategic support services. So, we also have an intranet that you can go to and get a policy available for the pack guideline. Then there are a lot of policies that structure and give you guidance on how to do HR policies” (CR1001=003, CW)*.

> *“There’s such a lot of policies available as a facility manager that you can go into the internet and get everything is there, and also SharePoint is available for documents” (CR1001=003, CW)*.

> *“We do have that system where we have all our policies on the internet where and you can scroll. Nobody needs to send you updates via the mail. You have an internal system that is connected to everybody in the whole Western Cape” (CR1001-00, CW)*.

Participants also noted that a key enabling factor was the ability to directly link the outcomes of the MDP to improvements in service delivery and operational challenges. For example, applying programme principles to solve issues such as inventory management allowed managers to see the immediate value of the training. This practical application of knowledge not only enhanced their problem-solving abilities but also reinforced the relevance of the programme within their day-to-day responsibilities, encouraging sustained engagement and buy-in.

> *“We support the district in terms of how they can improve, especially at the clinic level on how they can improve their inventory management, how to manage their stock, and how to take care of their assets” (CR1001-012, CW)*.

> *“They check the items in the store for expiring dates, so that we don’t throw those items away and have fruitful and wasteful expenditure…The ones that did the Aurum training, they still stick to the practice of standardizing on the products and to look at the quantity of items in their stores that store that they don’t waste money” (GR1001-002, GR)*.

> *“With the data, it seems like they understand the data better and with the quality improvement definitely because quality improved. The other thing is all these documents that the MDP is implementing or that can be implemented are also in line with the office of health standards” (GR1001-001, GR)*.

> *“The disaster plans. I mean, it’s one of the challenges that the subdistrict struggles with now that the MDP disaster plan tool is excellent” (GR1001-001, GR)*

However, participants also reported impactful operational constraints that limited their ability to fully implement and also engage with the MDP content. A key barrier was the systemic issue of **staff shortages**. Managers explained that the persistent understaffing at health facilities created a highly pressured environment with competing demands, which often left them with insufficient time and capacity to fully participate in training, mentorship, and the application of new skills. This operational impediment often mitigated full engagement with the programme, as managers were required to prioritize immediate service delivery needs and crisis management over strategic development and long-term planning.

> *“I do know there was maybe challenges to get the staff out of services due to staff shortages” (CR1001_004, CT)*

> *“The mentoring after the course the follow-up… it doesn’t have to be for a year but somebody that just go to these students where they’re working and support them with the implementation process… because with a shortage of staff and with everybody they’re so positive after completing the course and you don’t want them to lose. Yeah. if they can just a mentor to support them in practically, that will be excellent” (GR1001-001, GR)*.

> *“With a shortage of staff, they begin to shift services between the clinics. So, they will actually at one clinic they will do outreach services like physio and OT and that kind of stuff and then at the smaller clinics they will only do uh dispensing of chronic medicine” (GR1001-002, GR)*.

#### Prior exposure to PHC reforms e.g., Ideal Clinic Realisation and Maintenance (ICRM) implementation

Several participants highlighted that their prior involvement in national primary health care (PHC) reforms, such as the ICRM programme, contributed to creating an enabling environment for scaling the MDP by fostering integration and system-level thinking.

> *“So, the support is quite well and I think our substructure they always try to involve the other programs because of the holistic approach that we will, and because we work in the 2030 healthcare plan to make it a one-stop integrating service, especially with our ARVs and our women’s health” (CR1001-010, CT)*.

> *“We standardize the list and we also standardize on the type of equipment that we use in the clinics and the hospitals to make it easier for them, so that if they procure, they can procure centrally” (GR1001-002, GR)*.

While broader reforms, such as the National Health Insurance (NHI), were still in the early stages of implementation, initiatives like the Ideal Clinic had already laid important groundwork by fostering a shared understanding of performance standards.

> *“We have the Ideal clinic. So, I think IDEAL clinic has become and then obviously we know that the president approved the national health insurance act when that’s going to happen but yeah it was just a nice thing to approve it but implementation I have no idea when that’s going to happen” (CR1001-010, CT)*

Beyond the formal MDP curriculum, districts also reported the presence of pre-existing support structures for comprehensive health services that were well-aligned with the programme’s objectives. These structures, such as established mentoring networks and inter-disciplinary teams, provided a fertile ground for the application of skills learned in the MDP. For instance, participants were able to immediately apply problem-solving and team management techniques to their routine work within these existing frameworks, reinforcing the training and ensuring its practical relevance. This highlights that a key enabler for successful scalability is the ability of an intervention to leverage and integrate with a district’s established organizational ecosystem.

> *“The support structure for the PHC managers is that um we have a component um specifically, um for comprehensive health services and under comprehensive health services” (GR1001-003, GR)*.

#### Established training coordination units

Participants emphasized that established training coordination structures, such as the People Development Centre (PDC), play a critical role in supporting both the implementation and potential sustainability of the AURUM MDP. Centralized coordination through entities like the PDC was seen as essential for ensuring alignment with provincial priorities and promoting the long-term integration of the programme.

> *“All our training has been managed centrally by the People Development Centre (PDC), so that was where I linked mostly with AURUM. Because of this training for sustainability, we need to ensure that when AURUM exits that we can continue this program. So that is where we linked a prior career with PDC so that we can see how we can in the future, ensure that this is a sustainable program to continue with” (CT1001_004, CT)*.

Beyond centralized structures, participants highlighted that existing district and sub-district mechanisms are crucial for building capacity. These mechanisms, such as mentorship programmes, workshops, and team-based support, were identified as key components that complement formal training initiatives like the MDP. Respondents emphasized that integrating new leadership skills into routine practice is most effective when it is reinforced by an established support network. This finding indicates that for a programme to be truly scalable, it must be designed to leverage and strengthen these existing, decentralized support structures rather than operating as a standalone intervention.

> *“We work in an integrated way. so, for between district and sub-district there’s mentoring, there’s training, there’s meetings, there’s workshops, so on different levels to support the staff on ground level” (CT1001_004, CT)*.

Access to a variety of online and in-person courses, particularly those focusing on health systems management and finance, was also identified as an important factor in creating a supportive training environment. Participants noted that these additional training opportunities complemented the MDP by providing additional knowledge and reinforcing of key concepts. This supplementary education helped in building up a more comprehensive skillset, allowing managers to better understand the context in which they operate.

> *“There are also other online courses available that are specifically designed for the Department of Health and the systems that we use… There are also courses available for an introduction to finance management for non-financial managers. We provided access, there’s a budget for that” (CT1001_011, CT)*.

Finally, while participants acknowledged past training experiences, the AURUM MDP was often viewed as more practical and applicable to real-world district challenges: “

> *“So, for me, I did previously a management course, nursing management. We did cover some of the things or basically most of the things but in the MDP it was much more practical” (CT1001-001 001, CT)*.

### 2. Affordability and Resource Constraints

District managers described several financial and budgeting challenges that may influence the scalability and long-term sustainability of the MDP within the public health system. Participants consistently highlighted that budgeting processes are highly centralised and largely informed by historical expenditure patterns, leaving limited flexibility for introducing new leadership development initiatives. As one manager explained, *“It’s mainly historical budgeting, so what you spent last year really informs what the budget is for the next year” (CR1001-001, CW)*. While district and facility managers may contribute to budget discussions, participants perceived that final financial decision-making authority remains concentrated at provincial level, limiting local discretion to prioritise leadership development. Participants further described budgeting systems as rigid and heavily focused on core service delivery demands, particularly personnel expenditure, medicines, and consumables. One participant noted that “The budget for staff employment… is running at about between 70 and 80%” (GR1001-002, GR), leaving little flexibility for additional investments such as management training. Conditional grants were also described as restrictive, with one manager explaining *that “If you received a fund for HIV and TB, then I cannot fund training from that” (CR1001-008, CW)*. Within this constrained environment, leadership development initiatives were often perceived as competing with immediate operational and service delivery priorities.

Taken together, these findings suggest that the financial feasibility of scaling the MDP is influenced not only by available resources, but also by broader structural and governance factors within the public health system. Participants described constrained and highly regulated budgeting environments, limited district-level financial autonomy, and competing service delivery priorities as important barriers affecting sustained investment in leadership development initiatives. Despite these challenges, district managers consistently recognised the value of management training and its potential contribution to strengthening health system performance. As one participant reflected, *“The priority areas always take precedence, even when you know management training would help in the long run” (CR1001-009, CT)*. These accounts highlight the tension between immediate operational demands and longer-term investments in leadership development within district health systems.

### 3. District-Level Readiness and Support

In the context of scaling the MDP training, readiness refers to the extent to which a health system, particularly at the district level. It has the necessary structures, resources, leadership support, and organizational capacity in place to adopt, implement, and sustain the programme effectively.

Findings from this study revealed varied levels of district readiness and preparedness that could potentially impact scalability:

#### Procurement and Logistics Challenges

Participants consistently highlighted significant and systemic challenges in procurement and logistics, which negatively impact service delivery and resource management. A key concern was the frequent stockouts of essential medicines and consumables, which were attributed to weak procurement planning and inadequate forecasting. One participant explained:

> *“We really struggle with effective implementation; sometimes it’s resource constraints, sometimes it’s about…resistance to change”* (CR1001-001, CW).

Participants also pointed to limited control over tender processes and the complexity of upstream and downstream procurement decisions. As one manager explained:

> *“Sometimes you’re not directly in charge of the tenders,” and “there’s a lot of upstream and downstream factors…some of it is within your hands that you can change*” (CR1001-003, CW).

There was often a lack of coordination between supply chain staff and the teams providing services, which disrupted the smooth running of programmes. Participants explained that poor communication and unclear roles sometimes led to delays, duplication of tasks, or missed opportunities to apply what they had learned in the MDP training. It also makes it difficult for managers to fully apply the planning and problem-solving skills gained through the MDP.

> *“You’re dealing there with people who are not directly involved with the service…you’ll find that the priorities differ and the sense of urgency will be different”* (CR1001-006, CW).

Delays were made worse by all the paperwork and complicated admin processes, with participants describing how the heavy reliance on paper files slowed things down, created backlogs, and left both staff and clients feeling frustrated.:

> *“It’s paperwork and the system…then it’s to the supply chain, then only on the computer, then when you book it out”* (CR1001-005, CW).

#### Logistical Bottlenecks

Several logistical challenges were raised across all three districts, including transport difficulties and insufficient administrative support, all of which could affect the smooth implementation and potential scaling of the MDP. A common barrier was the lack of accountability and unclear roles in procurement processes, which led to delays:

> *“Employees don’t take responsibility… they don’t timely sign off documents,” and … There’s always a lack of understanding of what will be expected when someone is appointed*.*” (CR001-011, CT)*

In rural areas, geographic isolation posed additional constraints. One participant expressed frustration about the need to travel long distances for training:

> *“I don’t like to drive around to Cape Town for a week or two weeks for training*.*” (GR1001-002, GR)*

Additionally, the district has a constrained budget and limited flexibility. This highlights the financial limitations at the district level, where there are no dedicated funds for management training logistics. This gap is often addressed through reliance on donor-supported or externally funded programs, such as AURUM. One participant noted:

> *“All the controls that are put in place…are there for a reason, but it does result sometimes in a delay…limited budgets and budget cuts within a financial year”* (CR001-011, CT).

While monitoring and evaluation (M&E) are in place within the health system, they are often underutilised when it comes to tracking its outcomes. Participants highlighted that although trainings are delivered and new policies introduced, there is a limited follow-up to assess whether these initiatives translate into meaningful change in practice. One participant said:

> *“You can do training, have a new policy, you can do anything, but how do you know if things have changed? Are people implementing their new skills? Are they following through in terms of implementing the policy? I don’t think we do that effectively enough” (CR001-001, CW)*

### 4. Barriers to Scaling

Several key barriers to scaling the MDP emerged, highlighting challenges within the health system that limit a programme’s ability to expand beyond pilot to other districts or provinces. These include the costs and financial constraints of implementing the programme, human resource constraints, including staff shortages and staff resistance to change, as well as structural issues such as a lack of leadership support and limited integration of the programme into existing health system processes. A major barrier was the perception of the MDP’s cost in relation to its formal accreditation and return on investment.

> *“This is still work in progress to see how we can integrate this into our training of the province. And I think the concern regarding this training at this stage in its format is that if you compare value for money the NQF level is very low. and it’s a very expensive course”* (CR1001-004, CT).

Health managers frequently reported that day-to-day service delivery responsibilities limited their capacity to engage meaningfully with the programme. This means the routine demands such as staff supervision, patient care, managing procurements and responding to urgent operational challenges often took precedence over leadership development activities and skills gained through the programme:

> *“I must be honest with you. I battle to find the time to do the post-training work, which is very important because people have this training in the back of their heads but it’s not finding practical expression on a day-to-day basis” (CR1001-006, CW)*

> *“It addresses a lot of important issues but they don’t find time to do that because of human resource allocation. It was quite tough because there were a lot of assignments that they battled with. It was practical things that they could do to improve existing problems. But it does need follow-up because we are dealing here with people who have their fingers in a lot of pieces at one time” (CR1001-006, CW)*

Participants reported that resistance to change and difficulty shifting mindsets among some managers were seen as ongoing challenges to full adoption of MDP principles. While the programme introduced new ways of thinking about leadership, planning and accountability, participants reported that some managers are reluctant to to embrace adaptive leadership styles preferring old traditional approaches:

> *“Sometimes it takes three or four meetings just to get a mind change. Yes, it’s a change management thing. I can’t drive and I can’t ask and I can’t plead and I can’t beg and I can’t push you and I try to pull you. It is a very difficult situation there, I don’t want to say it’s not training that’s going to change them, training is not the solution for everything” (CR001-009, CW)*.

Participants noted that the impact of training was limited when the **post-training environments** did not provide opportunities to apply new skills. They highlighted that without supportive supervision, access to resources, the knowledge and skills gained during training were often underutilized, reducing the overall effectiveness of the MDP:

> *“If you can be trained in how to do the recruitment selection process but if nobody allows you to do it and give you the chance to develop in it for you, you will forget you need to practice certain things” (CR001-009, CW)*.

### 5. Facilitators and Opportunities

Several factors were identified as supportive of MDP scale-up, particularly those that facilitated integration, accessibility, and post-training continuity. Mentorship was consistently cited as a critical element to support implementation:

> *“I will say that the mentoring after the course, the follow-up. The mentoring…it doesn’t have to be for a year but somebody that just goes to these students where they’re working and supports them with the implementation process…because with a shortage of staff… Because it’s such a good course and I mean you actually want it to be implemented properly” (GR1001-001, GR)*.

Participants also suggested the programme’s long-term viability could be strengthened by embedding it within existing training structures. By integrating the programme into established systems rather than running it as a separate could reduce administrative and logistical costs for the government:

> *“I want to say that this training package needs to be linked with PDC to see how they can absorb it. That’s important for the province. So, whatever is the context and the way they are currently doing it PDC needs to absorb this and*…” (CR1001-004, CT).

Participants also mentioned that training offered within sub-districts will serve as a facilitator. This allowed more staff to participate as it minimized the need for long-distance travel and time away from routine duties. Participants noted that local delivery reduced logistical burdens such as arranging transportation, accommodation:

> *“What helps a lot is the fact that with the one training was within the sub-district, so we could send more people” (CR1001-005, CW)*.

Finally, strong advocacy from leadership and recognition of the programme’s value were seen as essential. Participants emphasized that when senior managers actively endorsed the programme, highlighted its benefits, and acknowledged staff achievements, it motivated wider engagement and fostered a culture of learning:

> *“Advocacy would be key because personally, I know that you would get much better gains. So that I think is something that we advocate on our side and advocacy from people like yourself who are actually hearing and understanding the impact that the program can actually assist us…” (CR1001-006, CW)*.

## Discussion

Findings from this study indicate that the AURUM MDP has inherent qualities that make it a highly scalable and adaptable intervention across different healthcare settings in South Africa (SA). The programme’s focus on fundamental leadership skills, including problem-solving, strategic planning, and effective team management, offers a universally applicable toolkit for middle and senior health managers [7]. These core competencies are not limited to a single operational environment but are relevant to a wide range of challenges faced within the public health system [3]. Findings from Orgill et al show how bottom-up leadership initiatives depend on existing structures, roles, and processes [11, 16].

The findings suggest that scalability is shaped less by the intervention itself and more by the contextual readiness of district health systems to absorb and sustain leadership development initiatives. Districts with stronger management systems, established accountability structures, supportive supervision, and existing cultures of organisational learning appeared better positioned to integrate the MDP into routine practice.

However, the programme’s adaptability is not uniform and is heavily influenced by the existing operational systems and support structures within a given district. Our analysis reveals that the effective integration of the MDP is contingent upon pre-existing management frameworks, established communication channels, and the presence of a supportive leadership culture. In districts with robust foundational systems, the MDP was more readily adopted and its principles were more easily embedded into routine practice. Conversely, in settings with weaker systems, programme implementation faced additional barriers, suggesting that successful scale-up requires a nuanced understanding of, and adaptation to, local context. This aligns with findings from mentorship and coaching interventions indicating that adapting to local leadership, having clear monitoring systems, and stakeholder engagement from the beginning are vital for sustainability and scale [17].

There was an indication that the strong support provided through the mentorship component of the MDP significantly enhanced their existing work structures. This finding suggests that the mentorship provided within the programme not only builds individual leadership skills but also serves as a catalyst for systemic improvements at the team and facility level. Similar systemic effects were observed in sub-Saharan Africa interventions where mentorship/coaching improved data use, leadership culture, and accountability in addition to clinical quality [17].

A prominent barrier was the systemic issue of staff shortages. This operational impediment often mitigated full engagement with the programme, as managers were required to prioritize immediate service delivery needs and crisis management over strategic development and long-term planning. These findings underscore that even a well-designed leadership programme can face implementation challenges when confronted with pre-existing, deeply rooted systemic issues. Evidence from SA shows that health worker shortages undermine quality service delivery, increase workloads, and reduce managers’ ability to engage in strategic tasks [18, 19].

The findings further highlight the importance of embedding leadership development within broader systems of continuous professional learning. Participants described how existing online platforms, mentoring structures, and district support systems strengthened implementation by reinforcing newly acquired skills and facilitating ongoing engagement beyond formal training sessions. These findings align with broader literature emphasizing that sustainable leadership development requires continuous learning environments rather than isolated training interventions [20, 21].

## Conclusion

The AURUM MDP demonstrates strong potential for scalability within South Africa’s public health system, particularly in districts with supportive leadership structures, established accountability systems, and existing organisational capacity for continuous learning. The findings highlight that successful scale-up depends not only on the programme itself, but also on the readiness of district health systems to integrate and sustain leadership development initiatives within routine practice. Mentorship, decentralised implementation approaches, and alignment with existing provincial training structures were identified as important enabling factors for long-term sustainability. However, operational pressures, workforce shortages, and constrained budgeting systems remain important barriers that may limit broader implementation. These findings provide important insights for future leadership development initiatives aimed at strengthening district health systems in low- and middle-income settings.

## List of abbreviations

ASLI: Appointment System Learnings Initiative
CT: Cape Town
CW: Cape Winelands
GCP: Good Clinical Practice
GR: Garden Route
HREC: Human Research Ethics Committee
ICH: Good Clinical Practice
ICRM: Ideal Clinic Realisation and Maintenance
ICH: International Council for Harmonisation’s
LMICs: Low- and middle-income countries
MDP: AURUM Management Development Programme
M&E: Monitoring and Evaluation
NHI: National Health Insurance
PDC: People Development Centre
PHC: Primary health care
SA: South Africa
UHC: Universal health coverage
WC: Western Cape
WHO: World Health Organization
ZAR: South African Rand

## Consent for publication

Not applicable

## Availability of data and materials

The datasets used and/or analysed during the current study are available from the corresponding author upon reasonable request.

## Competing interests

The authors declare no competing interests

## Funding statement

The Bill & Melinda Gates Foundation funded this research. INV-064789 Onoya (PI) 2024-2026. The contents herein are the responsibility of the authors and do not necessarily reflect the views of the Gates Foundation. Evaluation of the Aurum Management Development Programme in the Western Cape Province, South Africa

## Authors’ contributions

D. Onoya conceptualised the study, developed the research methodology, provided overall supervision and served as a principal investigator. C. Mongwenyana-Makhutle managed the study, collected the data, drafted the original manuscript and contributed to the interpretation of results. A. Moolla conducted the data analysis, contributed to the preparation of the manuscript and assisted with supervision. D. Hongoro performed the costing analysis and contributed to the drafting of the manuscript. T. Sineke, K. Shumba, and J. Miot contributed to the interpretation of results and provided a critical review of the manuscript. All authors reviewed and approved the final manuscript.

## Acknowledgments

We are particularly grateful to all the study participants for the time and information they shared with us. We thank the Western Cape Department of Health for their support and collaboration.

## Notes

### Competing Interest Statement

The authors have declared no competing interest.

